# Risk of perinatal deaths for fetuses and early newborns with congenital heart defects

**DOI:** 10.1101/2023.05.23.23290428

**Authors:** Xu Zhou, Yurong Jiang, Junqun Fang, Donghua Xie

## Abstract

**Background:** Few studies assessed the perinatal death (PD) risk for congenital heart defects (CHDs).

**Methods:** Data were obtained from the Birth Defects Surveillance System in Hunan Province, China, 2016–2020. Perinatal mortality (stillbirths and early neonatal deaths per 1000 fetuses (>=28 weeks)) and 95% confidence intervals (CI) were calculated. Crude odds ratios (ORs) and 95%CI were calculated to estimate the PD risk for CHDs and to examine the association of each maternal characteristic with PD risk for CHDs.

**Results:** 847755 fetuses were registered, including 14459 (1.71%) birth defects (4161 CHDs, 0.49%) and 833296 (98.29%) cases without birth defects. 7445 PDs (6874 stillbirths and 571 early neonatal deaths) were identified, including 976 (13.11%) CHD-related deaths, and 4396 PDs for fetuses without birth defects. 97.50% (936/960) of CHD-related stillbirths were selective termination of pregnancy. The overall perinatal mortality was 0.88% (95%CI: 0.86-0.90). CHDs increased the PD risk (23.46% vs. 0.53%, OR=57.78, 95%CI: 53.47-62.44). Any specific CHD increased the PD risk (OR>1, P<0.05). Perinatal mortality for CHDs was higher in rural than urban areas (OR= 3.40, 95%CI: 2.92-3.95). And perinatal mortalities for CHDs were higher in low maternal age (<25 years old), low per-capita annual income (<4000¥), low maternal education, no birth, and premature birth compared to the reference group. (OR>1, P<0.05).

**Conclusions:** CHDs significantly increase the PD risk. Several maternal characteristics were associated with CHD-related PDs. And several mechanisms have been proposed to explain these phenomena. Our results are of immediate value for clinical care and consultation.

## 1 Introduction

Perinatal deaths were the combination of fetal and neonatal deaths, including infant deaths that occur at less than 7 days of age and fetal deaths with a stated or presumed period of gestation of 28 weeks or more (1). Severe birth defects significantly increase the perinatal death risk (2-11). Congenital heart defect (CHD) is typically defined as a structural abnormality of the heart and/or great vessels present at birth (12). CHD is the most common birth defect (12-14), and about 20%–25% of specific CHDs were considered severe (15-17).

CHD is one of the most critical risk factors for children’s death, and most CHD-related deaths occur in low- and middle-income countries (18), which significantly burdens these countries. Previous studies have shown that CHDs increased the risk of adverse pregnancy outcomes, such as preterm birth, fetal brain damage and neonatal death (6, 19-25). And many studies regarded CHDs as adverse pregnancy outcomes (12, 13). However, few studies assessed the perinatal death risk for CHDs.

Notably, analysis of the epidemiology of perinatal death risk for CHDs has significant implications for clinical care and consultation. And it may contribute to reducing perinatal mortality.

Therefore, we conducted a comprehensive analysis based on the data from the Birth Defects Surveillance System in Hunan Province, south-central China (from 2016 to 2020), to define the relationship between CHDs and perinatal deaths. This study aimed to provide information on intervention programs to reduce perinatal mortality.

## 2 Methods

### 2.1 Data sources

This study used data from the Birth Defects Surveillance System in Hunan Province, China, 2016–2020, which is run by the Hunan Provincial Health Commission and involves 52 representative registered hospitals in Hunan Province. Surveillance data of fetuses (births and deaths at 28 weeks of gestation and beyond) and all birth defects included demographic characteristics such as residence, gender, maternal age, and other key information. Perinatal deaths include stillbirths (fetal deaths with gestation of 28 weeks or more) and early neonatal deaths (infant deaths less than 7 days of age). Perinatal mortality is the number of perinatal deaths per 1000 fetuses (births and deaths at 28 weeks of gestation and beyond).

Birth defects are diagnosed and classified according to the International Classification of Disease, Tenth Revision (ICD-10) (ICD codes: Q00-Q99), and ICD codes for CHDs were Q20-Q26.

To analyze the perinatal death risk of specific CHDs, if a fetus has more than one specific CHD, we consider only the major specific CHD according to the “Prognosis Grading and Perinatal Risk Assessment of Fetal Heart Disease” Expert Panel in China in 2022 (26).

### 2.2 Informed consents

We confirmed that informed consent was obtained from all subjects and/or their legal guardian(s). Doctors obtain consent from pregnant women before collecting surveillance data, witnessed by their families and the heads of the obstetrics or neonatal departments. Doctors obtain consent from their parents or guardians for live births, witnessed by their families and the heads of the obstetrics or neonatal departments. Since the Health Commission of Hunan Province collects those data, and the government has emphasized the privacy policy in the “Maternal and Child Health Monitoring Manual in Hunan Province”, there is no additional written informed consent.

### 2.3 Ethics guideline statement

The Medical Ethics Committee of Hunan Provincial Maternal and Child Health Care Hospital approved the study. (NO: 2022-S64). It is a retrospective study of medical records; all data were fully anonymized before we accessed them. Moreover, we de-identified the patient records before analysis. We confirmed that all experiments were performed following relevant guidelines and regulations.

### 2.4 Data quality control

To carry out surveillance, the Hunan Provincial Health Commission formulated the “Maternal and Child Health Monitoring Manual in Hunan Province”. Data were collected and reported by experienced doctors. To reduce integrity and information error rates of surveillance data, the Hunan Provincial Health Commission asked the technical guidance departments to conduct comprehensive quality control each year. For some complicated combined CHD, we ask experienced experts to identify it.

### 2.5 Materials and Data Availability

We had full access to all the data in the study and takes responsibility for its integrity and the data analysis. All data generated or analyzed during this study are included in this published article.

### 2.6 Statistical analysis

Using the log-binomial method, we calculated the perinatal mortality and its 95% confidence intervals (CI). And we calculated crude odds ratios (ORs) to examine the association between perinatal deaths and a broad range of birth defects.

Statistical analyses were performed using SPSS 18.0 (IBM Corp., NY, USA).

## 3 Results

### 3.1 Number of fetuses with or without CHDs

Our study included 847755 fetuses, including 14459 (1.71%) cases with birth defects and 833296 (98.29%) cases without birth defects. And 4161 (0.49%) cases with CHDs. **Table 1** shows the details of fetuses with or without birth defects by year. (**Table 1**)

**Table 1.**
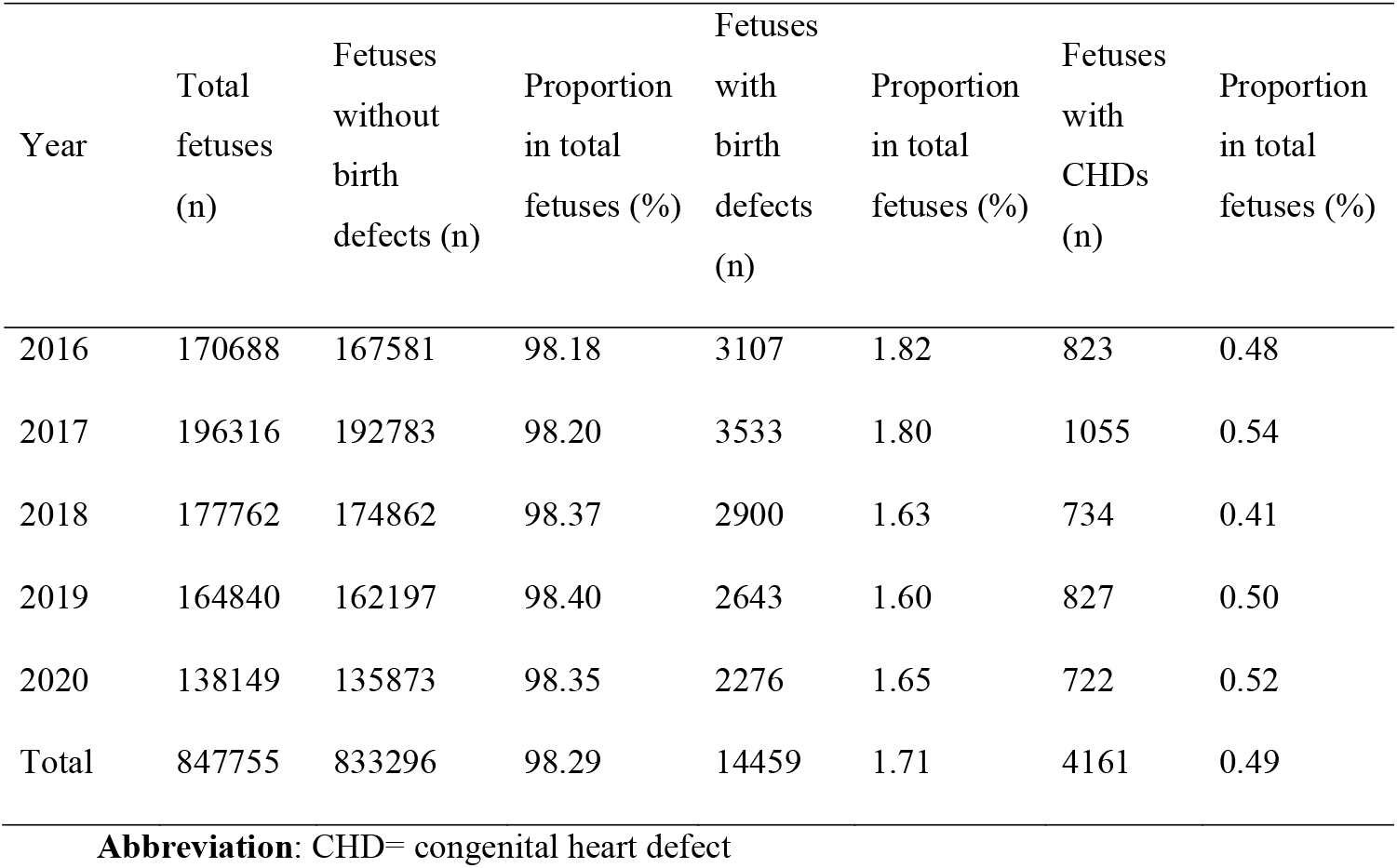
Number of fetuses with or without CHDs in Hunan Province, China, 2016-2020

| Year | Total<br>fetuses<br>(n) | Fetuses<br>without<br>birth<br>defects (n) | Proportion<br>in total<br>fetuses (%) | Fetuses<br>with<br>birth<br>defects<br>(n) | Proportion<br>in total<br>fetuses (%) | Fetuses<br>with<br>CHDs<br>(n) | Proportion<br>in total<br>fetuses (%) |
| --- | --- | --- | --- | --- | --- | --- | --- |
| 2016 | 170688 | 167581 | 98.18 | 3107 | 1.82 | 823 | 0.48 |
| 2017 | 196316 | 192783 | 98.20 | 3533 | 1.80 | 1055 | 0.54 |
| 2018 | 177762 | 174862 | 98.37 | 2900 | 1.63 | 734 | 0.41 |
| 2019 | 164840 | 162197 | 98.40 | 2643 | 1.60 | 827 | 0.50 |
| 2020 | 138149 | 135873 | 98.35 | 2276 | 1.65 | 722 | 0.52 |
| Total | 847755 | 833296 | 98.29 | 14459 | 1.71 | 4161 | 0.49 |

**Abbreviation:** CHD= congenital heart defect

### 3.2 CHD-related perinatal deaths

7445 perinatal deaths were identified, including 6874 stillbirths and 571 early neonatal deaths. 976 CHD-related deaths were identified, accounting for 13.11% of perinatal deaths. And 4396 perinatal deaths were related to fetuses without birth defects. 97.50% (936/960) of CHD-related stillbirths were selective termination of pregnancy. **Table 2** shows the details of CHD-related perinatal deaths by year. (**Table 2**)

**Table 2.**
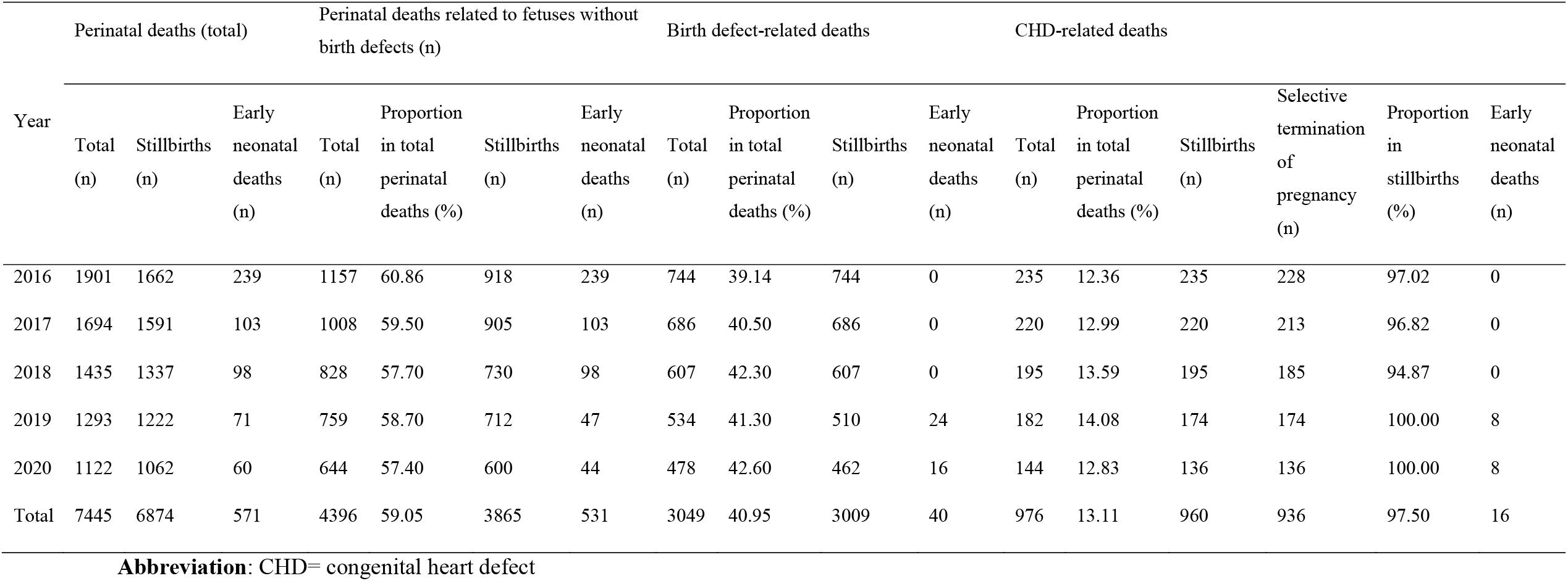
CHD-related perinatal deaths in Hunan Province, China, 2016-2020

### 3.3 Perinatal mortality and perinatal death risk for CHDs

The overall perinatal mortality was 0.88% (95%CI: 0.86-0.90). The perinatal mortality for fetuses with CHD was 23.46% (95%CI: 21.98,24.93), and perinatal mortality for fetuses without birth defects was 0.53% (95%CI: 0.51-0.54). Perinatal deaths were more common in fetuses with CHDs than fetuses without birth defects (OR=57.78, 95%CI: 53.47-62.44). **Table 3** shows the details of perinatal death risk for CHDs by year. (**Table 3**)

**Table 3.** Perinatal mortality and perinatal death risk for CHDs by year

| Year | Fetuses (total) |  |  | Fetuses (with CHDs) |  |  | Fetuses (without birth defects)<br>(Reference) |  |  | OR (95%CI) |
| --- | --- | --- | --- | --- | --- | --- | --- | --- | --- | --- |
|  | Total (n) | Perinatal deaths (n) | Perinatal mortality (%<br>95%CI) | Total (n) | Perinatal deaths (n) | Perinatal mortality (%<br>95%CI) | Total (n) | Perinatal deaths (n) | Perinatal mortality (%<br>95%CI) |  |
| 2016 | 170688 | 1901 | 1.11(1.06,1.16) | 823 | 235 | 28.55(24.90,32.20) | 167581 | 1157 | 0.69(0.65,0.73) | 57.49(48.89-67.59) |
| 2017 | 196316 | 1694 | 0.86(0.82,0.90) | 1055 | 220 | 20.85(18.10,23.61) | 192783 | 1008 | 0.52(0.49,0.56) | 50.13(42.68-58.88) |
| 2018 | 177762 | 1435 | 0.81(0.77,0.85) | 734 | 195 | 26.57(22.84,30.30) | 174862 | 828 | 0.47(0.44,0.51) | 76.04(63.68-90.81) |
| 2019 | 164840 | 1293 | 0.78(0.74,0.83) | 827 | 182 | 22.01(18.81,25.20) | 162197 | 759 | 0.47(0.43,0.50) | 60.02(50.17-71.80) |
| 2020 | 138149 | 1122 | 0.81(0.76,0.86) | 722 | 144 | 19.94(16.69,23.20) | 135873 | 644 | 0.47(0.44,0.51) | 52.31(42.90-63.79) |
| Total | 847755 | 7445 | 0.88(0.86,0.90) | 4161 | 976 | 23.46(21.98,24.93) | 833296 | 4396 | 0.53(0.51,0.54) | 57.78(53.47-62.44) |

**Abbreviation:** CHD= congenital heart defect; CI= confidence intervals; OR= crude odds ratio

### 3.4 Perinatal mortality and perinatal death risk for specific CHDs

Any specific CHD increased the perinatal death risk (OR>1, P<0.05). Ventricular septal defect (OR= 20.31, 95%CI: 17.66-23.35), atrial septal defect (OR= 5.31, 95%CI: 3.12-9.04), and Tetralogy of Fallot (OR= 397.17, 95%CI: 280.32-562.75) were the most common specific CHDs. And Tetralogy of Fallot (OR= 397.17, 95%CI: 280.32-562.75) and double outlet right ventricle (OR= 387.04, 95%CI: 223.49-670.27) were with the highest perinatal death risk. **Table 4** shows the details of perinatal mortality and perinatal death risk for specific CHDs. (**Table 4**)

**Table 4.**
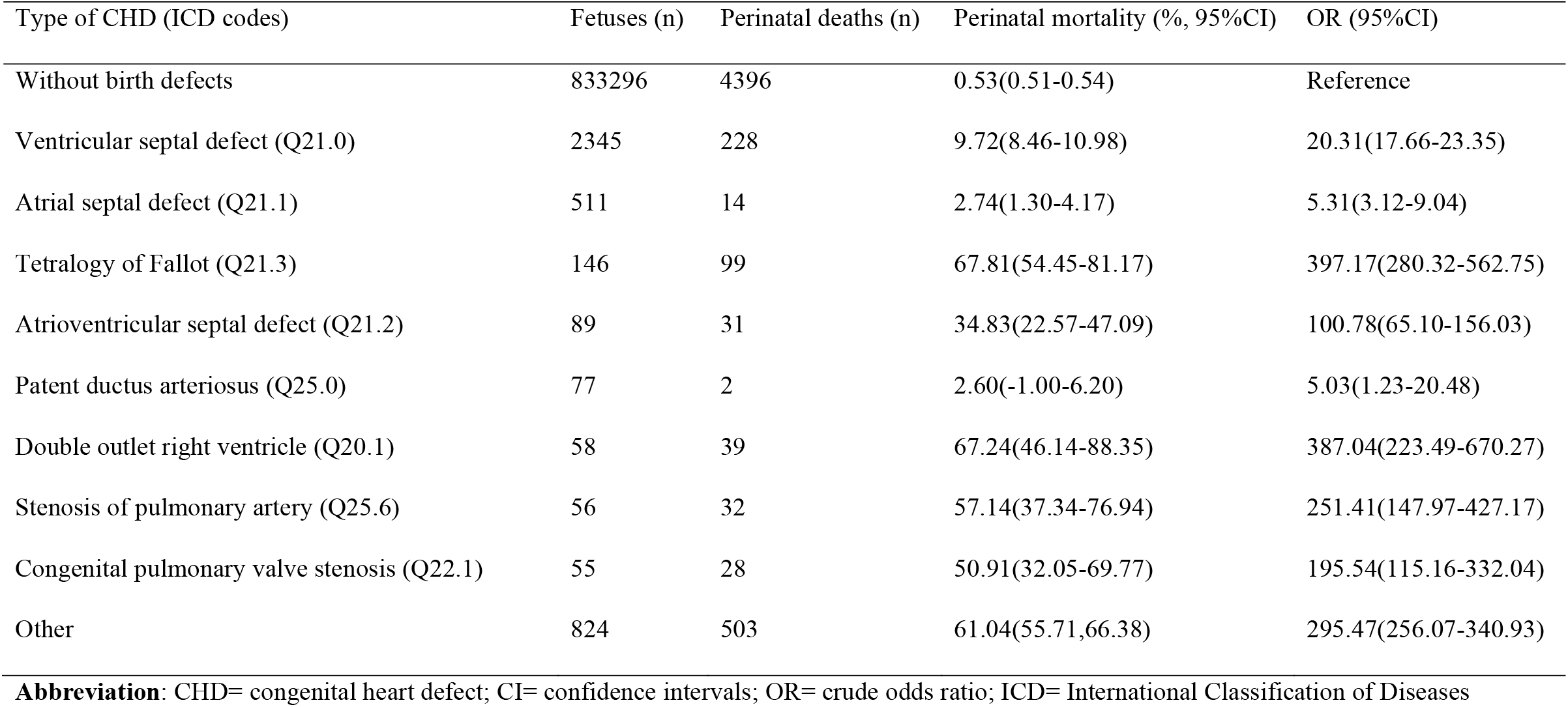
Perinatal mortality and perinatal death risk for specific CHDs

| Type of CHD (ICD codes) | Fetuses (n) | Perinatal deaths (n) | Perinatal mortality (%; 95%CI) | OR (95%CI) |
| --- | --- | --- | --- | --- |
| Without birth defects | 833296 | 4396 | 0.53(0.51-0.54) | Reference |
| Ventricular septal defect (Q21.0) | 2345 | 228 | 9.72(8.46-10.98) | 20.31(17.66-23.35) |
| Atrial septal defect (Q21.1) | 511 | 14 | 2.74(1.30-4.17) | 5.31(3.12-9.04) |
| Tetralogy of Fallot (Q21.3) | 146 | 99 | 67.81(54.45-81.17) | 397.17(280.32-562.75) |
| Atrioventricular septal defect (Q21.2) | 89 | 31 | 34.83(22.57-47.09) | 100.78(65.10-156.03) |
| Patent ductus arteriosus (Q25.0) | 77 | 2 | 2.60(-1.00-6.20) | 5.03(1.23-20.48) |
| Double outlet right ventricle (Q20.1) | 58 | 39 | 67.24(46.14-88.35) | 387.04(223.49-670.27) |
| Stenosis of pulmonary artery (Q25.6) | 56 | 32 | 57.14(37.34-76.94) | 251.41(147.97-427.17) |
| Congenital pulmonary valve stenosis (Q22.1) | 55 | 28 | 50.91(32.05-69.77) | 195.54(115.16-332.04) |
| Other | 824 | 503 | 61.04(55.71,66.38) | 295.47(256.07-340.93) |

**Abbreviation:** CHD= congenital heart defect; CI= confidence intervals; OR= crude odds ratio; ICD= International Classification of Diseases

### 3.5 Epidemiology of perinatal death risk for CHDs

Perinatal mortality for CHDs was higher in rural than urban areas (OR= 3.40, 95%CI: 2.92-3.95). Compared to the reference group, perinatal mortalities for CHDs were higher in the following group (OR>1, P<0.05): low maternal age (<25 years old), low per-capita annual income (<4000¥), low maternal education, no birth, and premature birth. **Table 5** shows the details of perinatal death risk for CHDs by epidemiological characteristics. (**Table 5**).

**Table 5.** Epidemiology of perinatal mortality and perinatal death risk for CHDs

| Characteristics | Fetuses<br>with CHDs<br>(n) | Perinatal<br>deaths<br>(n) | Perinatal mortality<br>(%, 95%CI) | OR (95%CI) |
| --- | --- | --- | --- | --- |
| Region |  |  |  |  |
| Urban | 2342 | 328 | 14.01(12.49-15.52) | Reference |
| Rural | 1819 | 648 | 35.62(32.88-38.37) | 3.40(2.92-3.95) |
| Gender |  |  |  |  |
| Male | 2204 | 514 | 23.32(21.31-25.34) | Reference |
| Female | 1952 | 457 | 23.41(21.27-25.56) | 1.01(0.87-1.16) |
| Unknown | 5 | 5 | 100.00(12.35-187.65) | - |
| Maternal age (years old) |  |  |  |  |
| <20 | 69 | 32 | 46.38(30.31-62.45) | 2.92(1.80-4.76) |
| 20-24 | 474 | 176 | 37.13(31.65-42.62) | 2.00(1.61-2.48) |
| 25-29 | 1700 | 388 | 22.82(20.55-25.09) | Reference |
| 30-34 | 1270 | 248 | 19.53(17.10-21.96) | 0.82(0.69-0.98) |
| >=35 | 648 | 132 | 20.37(16.90-23.85) | 0.87(0.69-1.08) |
| Per-capita annual income (¥) |  |  |  |  |
| <2000 | 36 | 17 | 47.22(24.77-69.67) | 2.79(1.44-5.40) |
| 2000- | 367 | 127 | 34.60(28.59-40.62) | 1.65(1.31-2.08) |
| 4000- | 1235 | 219 | 17.73(15.38-20.08) | 0.67(0.57-0.80) |
| 8000- | 2523 | 613 | 24.30(22.37-26.22) | Reference |
| Maternal education |  |  |  |  |
| Illiteracy or primary school | 50 | 20 | 40.00(22.47-57.53) | 1.93(1.08-3.43) |
| Secondary school | 537 | 181 | 33.71(28.80-38.62) | 1.47(1.19-1.82) |
| Senior school | 1540 | 396 | 25.71(23.18-28.25) | Reference |
| University or above | 2034 | 379 | 18.63(16.76-20.51) | 0.66(0.56-0.78) |
| Number of births |  |  |  |  |
| 0 | 38 | 30 | 78.95(50.70-107.20) | 12.62(5.75-27.72) |
| 1 | 2122 | 486 | 22.90(20.87-24.94) | Reference |
| 2 | 1756 | 397 | 22.61(20.38-24.83) | 0.98(0.85-1.14) |
| ≥3 | 245 | 63 | 25.71(19.36-32.06) | 1.17(0.86-1.58) |
| Gestational age of termination |  |  |  |  |
| <34 weeks | 910 | 701 | 77.03(71.33-82.74) | 77.59(60.48-99.53) |
| 34-36 weeks | 717 | 170 | 23.71(20.15-27.27) | 7.19(5.54-9.33) |
| ≥37 weeks | 2534 | 105 | 4.14(3.35-4.94) | Reference |

**Abbreviation:** CHD= congenital heart defect; CI= confidence intervals; OR= crude odds ratio

## 4 Discussion

Overall, perinatal mortality for CHDs was relatively high. CHDs (including any specific CHD) significantly increased the perinatal death risk. And CHD-related perinatal deaths were more common in rural areas, mothers with low maternal age, low per-capita annual income, low maternal education, no birth, and premature birth. Our study is the first to systematically analyze the perinatal death risk for CHDs. Our results are of immediate value for clinical care and consultation.

CHDs significantly increased the perinatal death risk, and most CHD-related perinatal deaths were selective terminations of pregnancy. Previous studies showed birth defects increased perinatal death risk (2, 8, 27, 28). It is similar to ours. The perinatal mortality for CHDs (23.46%) was higher than the overall perinatal mortality for birth defects. E.g., Elisabeth et al. reported that 14% (1563/11353) of pregnancies with birth defects were ended by a second-trimester termination (29). Dominique et al. reported that 852 were stillborn in 19170 eligible neonates and fetuses with birth defects (4.44%) (8). We also found any specific CHD increased the perinatal death risk. It is similar to previous studies (30, 31). And Tetralogy of Fallot and double outlet right ventricle had the highest perinatal death risk. It indicates that these specific CHDs are relatively more severe (24, 32-37). Our study is the first to systematically give the perinatal mortality and perinatal death risk (OR values) for specific CHDs.

We also found that CHD-related perinatal deaths were more common in rural areas, mothers with low maternal age, low per-capita annual income, low maternal education, no birth, and premature birth. And there are several mechanisms to explain these phenomena. First, low maternal age and no birth were associated indicators. Younger women are more likely to terminate a pregnancy with CHD because they can get pregnant again easily. In contrast, women of advanced age have difficulty conceiving and are more likely to keep fetuses with birth defects (38). Second, women in rural areas, low per-capita annual income, and low maternal education were also associated indicators. It may be mainly related to medical conditions and economic conditions. Better medical conditions and economic conditions benefit children with CHD surviving (39, 40). Third, as most CHD-related deaths were selective termination of pregnancy, the earlier pregnancy with birth defects is terminated, the less impact it will have on the pregnant woman (41, 42). It is the reason that perinatal deaths are more common in premature birth.

Some things could be improved in our study. E.g., many fetuses have more than one birth defect, but we do not analyze their combined effect.

In summary, perinatal mortality for CHDs was relatively high, and CHDs significantly increased the perinatal death risk. CHD-related perinatal deaths were more common in rural areas, mothers with low maternal age, low per-capita annual income, low maternal education, no birth, and premature birth. And several mechanisms have been proposed to explain these phenomena. Our results are of immediate value for clinical care and consultation.

## Data Availability

All data generated or analyzed during this study are included in this published article.

## 5 Acknowledgments

The authors thank the staff working for the Birth Defects Surveillance System of Hunan Province, China, from 2016 to 2020.

## 8 Supplemental Material

Ethical approval.pdf

